# Work-Related Musculoskeletal Disorders among Nurses Working at Hospitals of Sudurpaschim Province, Nepal

**DOI:** 10.1101/2023.09.24.23296044

**Authors:** Deepa Kumari Bhatta, Gajananda Prakash Bhandari, Nirmal Duwadi, Bal Krishna Bhatta, Ishwori Gurung, Alisha Dahal

**Affiliations:** Department of Public Health, Nobel College, Pokhara University, Kathmandu, Nepal; Executive Member, Prevent Cancer Nepal

**Author notes:** These authors contributed equally to this work (Deepa Kumari Bhatta, Gajananda Prakash Bhandari, Nirmal Duwadi). These authors also contributed equally to this work (Ishwori Gurung, Alisha Dahal).

**Keywords:** Works related to musculoskeletal disorders, nursing, prevention, perceived risk factors

## Abstract

Musculoskeletal conditions have been ranked as the leading cause of disability worldwide. Low back pain is the single largest contributor in 160 countries including Nepal. Nurses working in hospitals in Nepal are overworked and at risk of developing musculoskeletal disorders. The study aimed to determine the 12-month periods and point prevalence of work-related musculoskeletal disorders (WMSDs); the predictors, and the perceived risk factors among nurses. A self-administered standard Nordic questionnaire was distributed among all eligible 118 nurses working at two hospitals. Bivariate and multivariate logistic regression analyses were applied to identify the predictors of WMSDs. Study findings revealed that nearly half (47.46%) of nurses had WMSDs most significantly affecting the lower back (18.93%). Nurses exceeding 30 years of age had almost seven times higher odds of having WMSDs compared to their counterparts aged less than 30 adjusting the effects of BMI, department, years of clinical experience, and type of hospital (OR= 6.92; CI =1.67-28.58). There was decreased odds of experiencing WMSDs by 0.33 times for nurses working in a private hospital than in a government hospital adjusting the effect of age, BMI, department, and years of clinical experience (OR=0.33; 95% CI=0.12-0.91). Similarly, nurses working in critical units had 5.24 times higher odds of having musculoskeletal disorders than in general units (OR=5.25; 95% CI=1.79-15.38). Nurses working for more than five years had 7.53 times higher odds of having WMSDs than those with less than and equal to five years of work experience (OR=7.53; CI 2.78-20.32). WMSDs are common among nurses primarily affecting the lower back and the odds of having MSDs is high with increasing age, BMI, and work experience, and for nurses who worked in critical care units and public hospital. Prompt preventive measures should be adopted to avoid risk factors of MSDs in work settings. **Keywords:** Works related to musculoskeletal disorders, nursing, prevention, perceived risk factors

## Introduction

Musculoskeletal disorders (MSDs) are painful degenerative and inflammatory conditions. The symptoms include stiff joints, dull aches, swelling, and recurrent pain (1,2). Musculoskeletal conditions are the most common causes of disability and limitation related to daily living and gainful employment (3).

MSDs are considered a “new epidemic” disease by the International Labour Organization (ILO) and the World Health Organization (WHO) that needs to be researched and solved. MSDs significantly impact work-related absence, accounting for a large proportion of missed days (4). The Global Burden of Disease (GBD 2019) has indicated low back pain (LBP) as the leading cause of disability associated with a significant amount of cost (5–7). Approximately 2.35 million people live with MSDs in Nepal where low back and neck pain has been ranked as the leading cause of disability in Nepal with a 16.9% increase in prevalence from 2005 to 2016 (8,9). A high-risk group for MSDs has been identified as nurses (10). Studies have shown that the prevalence rate of MSDs among nurses is usually high ranging between 60-95% (11,12). Low back pain (LBP) is the most commonly reported MSD among nurses, with prevalence ranging from 33 to 90.1% worldwide (5,13). A variety of factors have been linked to the development of MSDs□including physical factors such as patient handling and long-standing hours□psychosocial factors such as stress, anxiety, and depression□and organizational factors such as working shifts, staff shortages, and poor working conditions (14,15). As a result WMSDs pose a significant threat to the quality of life of nurses□resulting in work absenteeism, work limitations, and the eventual need to change jobs (16).

The nursing profession is a very demanding job, both physically and emotionally making nurses more susceptible to work-related musculoskeletal disorders (WMSDs). Besides the physical demands of the job, the working hours increases the likelihood of WMSDs (17,18). Hospital nurses in Nepal are overworked because of the higher nurse-patient ratio. In Nepal, nurses are therefore expected to have a substantially high risk of getting musculoskeletal illnesses and discomfort associated with them (17). In a study conducted among more than 500 nurses working in different departments at Tribhuvan University Teaching Hospital, Kathmandu, Nepal in 2019, the prevalence of low back pain was 64.5% (19). Working under physical overload due to long work hours and patient handling demands leads to a high risk of developing WMSDs hampers the working efficiency of nurses, and has a direct impact on patient safety in clinical practice (20,21).

Although the nursing profession is known to be a high risk for WMSDs, to the best of our knowledge, it is one of the least-studied occupations in Nepal (19). Determining the prevalence and factors associated with WMSDs among nurses as identified by problem analysis S1 Fig is important for hospital administrators, managers, healthcare workers, and health policymakers to reduce musculoskeletal discomfort among nurses and thereby achieve improved nursing performance. Thus, this study aimed to assess the burden of MSDs and possible risk factors among nurses working at different hospitals in Sudurpaschim Province.

## Material and Methods

### Study design

The study employed a descriptive analytical cross-sectional design which used a self-administered questionnaire.

### Study site

The research was carried out at two hospitals in Nepal’s Sudurpaschim Province. (1) Mahakali Provincial Hospital, which is a provincial government-owned hospital, and (2) Nisarga Hospital and Research Center, which is a privately owned hospital. These hospitals provide services at both the regional and district levels. These hospitals serve a diverse range of patients from various socioeconomic backgrounds and communities.

### Inclusion and exclusion criteria

Survey questionnaires were distributed to all eligible nurses with at least 1 year of work experience.

Nurses who were pregnant were on study and maternity leave, who had suffered recent trauma injury or serious diseases such as diabetes, disc prolapse, and rheumatoid arthritis, and part-time worker were excluded from the study.

### Sample size

Based on published material by (17), the prevalence of work-related musculoskeletal disorders (WRMSDs) among nurses of Kaski, Nepal was reported as being 86% (0.86). Using a statistical significance α of 0.05, precision at 0.05, and estimated prevalence of WRMSDs as 0.86 for an unknown population size, 185 nurses was the required sample size. The sample size estimated using a finite population is 102. However, all 118 of the nurses working at both hospitals meeting inclusion criteria were included in the study.

### Study period and Data collection

The study period was from April to July 2022. Questionnaires were distributed by a researcher with verbal instruction to each nurse who was on duty after taking hospital authority consent. The name list of all working nurses was prepared coordinating with the nursing administrator. The checklist was prepared to keep track record of a questionnaire distributed and responses collected. A daily follow-up visit was done to collect the responses and as a gentle reminder to fill up the questionnaires.

### Study Tool

Self-administered questionnaire S1 File in Nepali language including four parts and a patient information sheet with an informed consent form was used. The first part of the questionnaire includes questions related to the demographic and work-related characteristics of the nurses. The second part of the tool was adapted from the Nordic Musculoskeletal Questionnaire and consists of questions referring to nine body areas. The respondents were asked to select from nine body locations (shoulder, neck, lower back, upper back, elbow joint, hand or wrist, hip joint, knee joint, and ankle joint). The questions include whether they have felt discomfort in the past year. This questionnaire was designed as a standard questionnaire and has adequate internal consistency, reliability, and validity (22). The third part of the questionnaire includes the measures taken in response to the MSD experienced by the nurses. The fourth part of the questionnaire consists of questions on the perception of nurses on factors that may contribute to the development of musculoskeletal disorders among nurses. It included 17 conditions and tasks at work that could contribute to musculoskeletal disorders to be scored on a scale of 0 to 10 (14).

### Ethical consideration

This study was approved by the Ethical Review Committee of the Nobel College (Reg. No. MPHIR0027/2022) S2 Fig. The study’s objectives and the questionnaire were explained to all participants, and they were assured of their anonymity and the confidentiality of their responses. To participate, all participants signed an informed consent form.

## Data analyses

Data was analyzed using the IBM SPSS software statistical version 16.0 for Windows (IBM, Armonk, NY, USA). The proportion of MSDs was summarized using the descriptive statistics of mean, confidence interval, and percentages. Chi-square analysis was used to determine the association of self-reported musculoskeletal symptoms with socio-demographic, personal, and work-related characteristics of nurses. Logistic regressions were used to determine risk factors related to musculoskeletal discomfort. Adjusted and unadjusted odds ratios (OR) and upper and lower 95% confidence intervals (CI) were calculated to determine the contribution of each factor. The significant α level was set at 0.05.

## Results

The descriptive information of all respondents is presented in Table 1. The mean age, height, weight, body mass index, and work experiences of the respondents were 27.91 years, CI 26.56-29.2636.4; 1.56 meter CI 1.55-1.57; 56.63 kg, CI 54.95-58.32; 23.37 kg/m^2^, CI 22.57-24.17 and 5.45 years, CI 4.39-6.51 respectively. Nearly three fifth (58.47%) of the respondents were married followed by single (40.67%) and divorced (0.84%). More than half of the respondents (56.78%) reported to engage in physical exercise. Among them, nearly three-fifths (59.70%) of the respondents said they exercise daily. Almost all (99.15%) of the respondents were non-smokers. Slightly more than half (51.69%) of the respondents were working in private hospital and the remaining 48.31 percent of the respondents were working in government hospital. More than half (56.78%) of the respondents were working in general wards followed by slightly more than a quarter (26.27%) working in the critical unit, 8.47 percent in OT, 6.78 percent in the postoperative ward, and least were working in emergency wards (1.69%). All respondents reported working six days a week, with one day off, for a total of more than 40 hours per week.

**Table 1.**
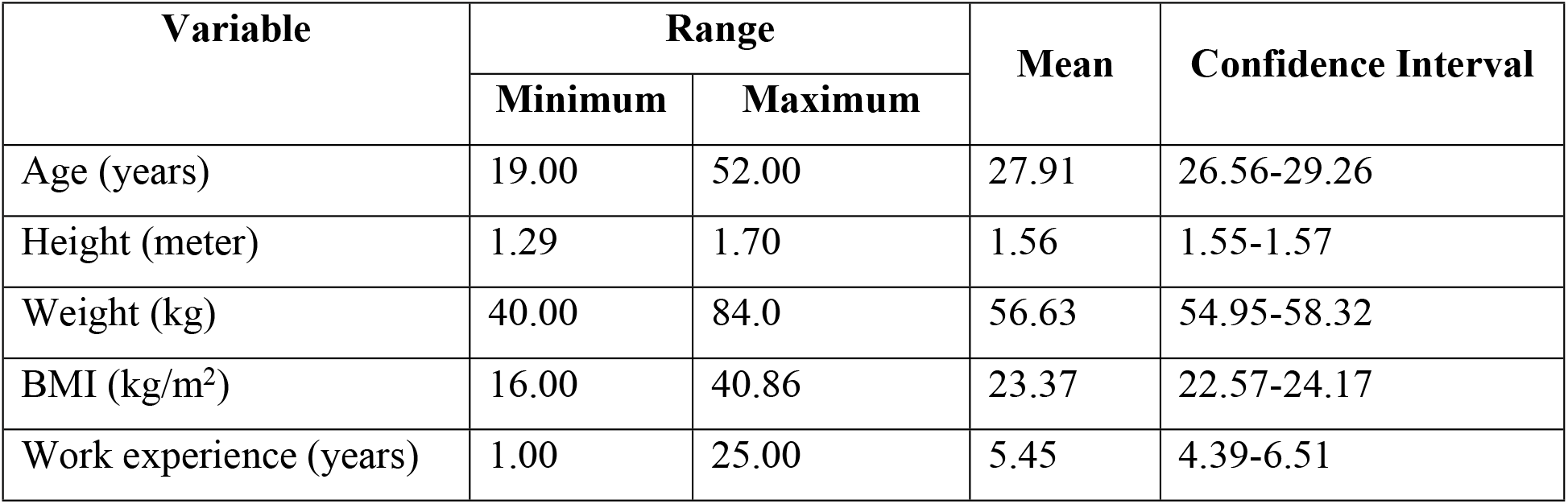
Descriptive information of all respondents.

Of the respondents, nearly half (47.46%) of the respondents reported musculoskeletal disorders persisting in at least one of the 9 anatomical sites and lasting for more than 3 days in the previous 12 months. Fig 1 show**s** that the 12-month prevalence rates of WMSDs was highest in the lower back (57.14%), followed by the feet/ankles (46.43%) and then shoulder (35.71%) but least in the elbow (10.71%).

**Fig 1.**
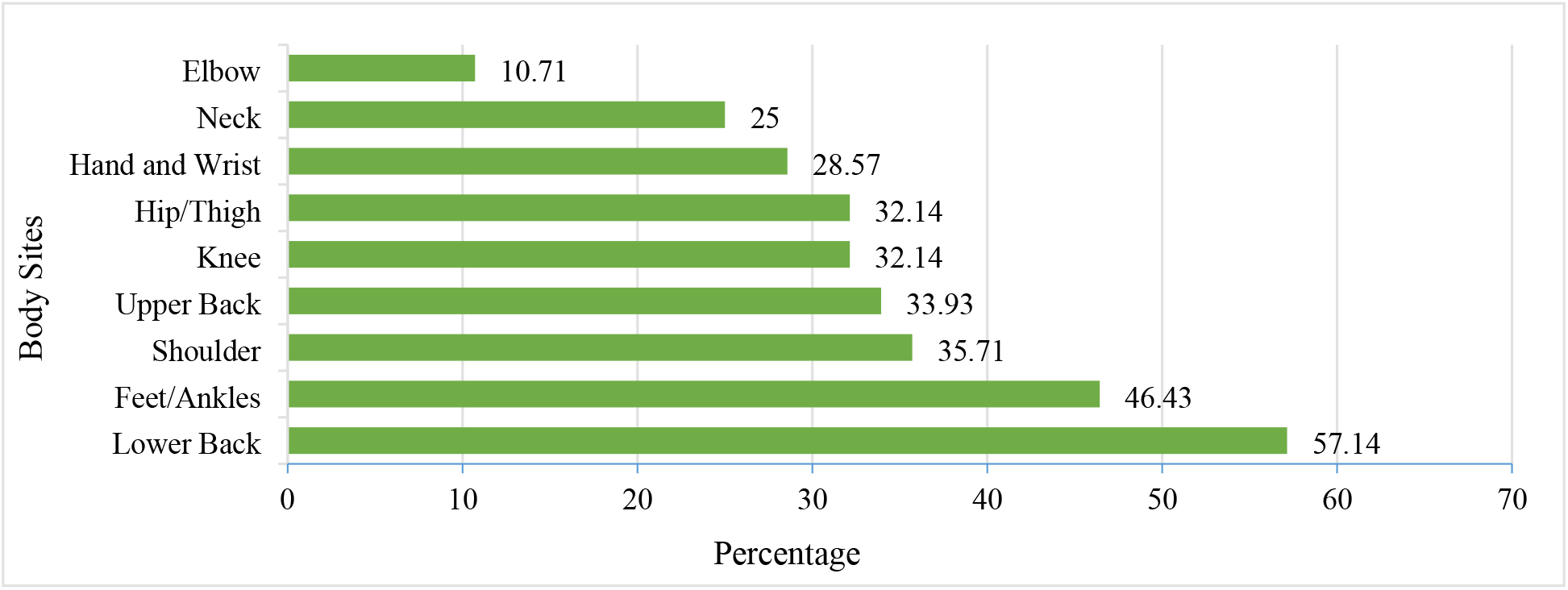
Proportion of work-related musculoskeletal disorders for different body sites among nurses.

Of all the respondents that indicated WMSDs only 51.79% reported that they had sought treatment for WMSDs. followed by 48.21 percent who said that they were involved in exercise and posture programs after experiencing musculoskeletal disorders. Similarly, 46.43 percent of respondents took sick leave, followed by 44.64 percent who took prescribed medication for musculoskeletal disorders. Similar least percentage (10.71%) of the respondents changed duties and changed type of patient treating respectively due to MSDs. The lowest percentage (3.57%) of the respondents decreased patient contact hours due to MSDs Table 2.

**Table 2.** Percentage indicating measures followed by respondents toward prevention and control of work-related musculoskeletal disorders.

| Measures/Effects | Percentage |
| --- | --- |
| Sought treatment or consultation | 51.79 |
| Exercise or posture program | 48.21 |
| Took time off on sick leave | 46.43 |
| Took prescribed medication | 44.64 |
| Modified techniques and procedures | 32.14 |
| Sought alternative treatments | 16.07 |
| Changed work setting | 12.50 |
| Changed duties | 10.71 |
| Changed type of patient predominantly treating | 10.71 |
| Decreased patient contact hours | 3.57 |

Table 3 includes the results of the perception of respondents towards the various work-related 17 conditions and tasks that can contribute to the development of MSDs. The perception for each condition was measured with a 1 to 10 score rating scale. A score of greater than 7 indicated that a condition was considered a **major risk factor**. Table 3 shows the respondents’ perceptions of the risk factors that could contribute to work-related discomfort and injury. The respondents who had experienced WMSDs indicated that working in the same positions for long periods (69.64%), reaching or working away from your body (67.86%) and working schedule (64.29%) were the most perceived job risk factors precipitating WMSDs during their clinical practice.

**Table 3.** Perceived risk factors contributing to the development of WMSDs.

| **Work-related risk factors\*** | **Number** | **Percentage** |
| --- | --- | --- |
| Working in the same position for long periods | 39 | 69.64 |
| Reaching or working away from your body | 38 | 67.86 |
| Work scheduling (Overtime, Irregular Shift, etc.) | 36 | 64.29 |
| Inadequate training on injury prevention | 33 | 58.93 |
| Not enough rest breaks during the workday | 31 | 55.36 |
| Working in an awkward or cramped position | 29 | 51.79 |
| Attending an excessive number of patients during day | 29 | 51.79 |
| Continuing to work while injured or hurt | 26 | 46.43 |
| Lifting or transferring dependent patient | 26 | 46.43 |
| Working with confused and agitated patients | 25 | 44.64 |
| Unanticipated sudden movement or fall of the patient | 24 | 42.86 |
| Performing the same task over and over | 21 | 37.50 |
| Bending or twisting your back in an awkward way | 20 | 35.71 |
| Carrying, lifting, or moving materials or equipment | 17 | 30.36 |
| Performing manual nursing techniques | 9 | 16.07 |
| Assisting patients during gait activities | 9 | 16.07 |

**Table 4.** Association of self-reported 12-month prevalence of work-related musculoskeletal disorders, and demographic and personal factors of all the respondents.

| Characteristics | MSDs n(%) |  | X<sup>2</sup> | p-value |
| --- | --- | --- | --- | --- |
|  | Yes | No |  |  |
| **Age (In years)** |  |  |  |  |
| ≤30 | 30 (53.57) | 57 (91.94) | 22.36 | **0.001\*** |
| >30 | 26 (46.43) | 5 (8.06) |  |  |
| **Marital Status** |  |  |  |  |
| Single | 20 (35.71) | 29 (46.77) | 1.48 | 0.22 |
| Married | 36 (64.29) | 33 (53.23) |  |  |
| **BMI** |  |  |  |  |
| Underweight | 2 (3.57) | 6 (9.68) | 8.53 | **0.01\*** |
| Normal weight | 31 (55.36) | 45 (72.58) |  |  |
| Overweight/Obesity | 23 (41.07) | 11 (17.74) |  |  |
| **Physical Exercise** |  |  |  |  |
| Yes | 27 (48.21) | 24 (38.71) | 1.08 | 0.29 |
| No | 29 (51.79) | 38 (61.29) |  |  |
| **Type of Hospital** |  |  |  |  |
| Government | 39 (69.64) | 18 (29.03) | 19.43 | **0.001\*** |
| Private | 17 (30.36) | 44 (70.97) |  |  |
| **Working department** |  |  |  |  |
| General wards | 25 (44.64) | 42 (67.74) | 6.39 | **0.01\*** |
| Critical units | 31 (55.36) | 20 (32.26) |  |  |
| **Years of clinical experience** |  |  |  |  |
| ≤5 years | 31 (35.63) | 56 (64.37) | 16.81 | **0.001\*** |
| >5 years | 25 (80.65) | 6 (19.35) |  |  |

**\*p-value<0.05**

Further finding the risk factors that could contribute to MSDs among nurses suggested that nurses with age greater than 30 years had 6.92 times higher odds of having musculoskeletal disorders than nurses with age less than 30 years (AOR= 6.92; CI =1.67-28.58). The odd of experiencing MSDs is decreased by 0.33 times for nurses working in a private hospital than nurses working in government (AOR=0.33 CI=0.12-0.91). The unadjusted odds ratio of musculoskeletal disorders for overweight and obese nurses is 3.04 times higher than nurses with normal weight (COR 3.04; 95% CI 1.29-7.11). However, there is no effect of BMI on WMSD after adjusting the effect of other explanatory variables. Nurses working in critical units had 5.24 times higher odds of musculoskeletal disorders than nurses working in general units (AOR=5.24 CI=1.79-15.38). The odds of musculoskeletal disorders increase by 7.53 times for nurses with work experience of more than five years than for nurses with less than five years of work experience (COR=7.53; CI 2.78-20.32). While adjusting other explanatory variables there is no effect of years of clinical experience on WMSD (Table 5).

**Table 5.** Risk factors for WMSDs.

| Factors | MSD<br>n (%) | Unadjusted OR<br>(95% CI) | p-value | Adjusted OR<br>(95% CI) | p-value |
| --- | --- | --- | --- | --- | --- |
| **Age (years)** |  |  |  |  |  |
| >30 | 26 | 9.88 (3.44-28.35) | **<0.001\*** | 6.92 (1.67-28.58) | **0.007\*** |
| **Type of Hospital** |  |  |  |  |  |
| Private | 39 | 0.18 (0.08-0.39) | **<0.001\*** | 0.33 (0.12-0.91) | **0.031\*** |
| **BMI** |  |  |  |  |  |
| Underweight | 2 | 0.48 (0.09-2.56) | 0.39 | 1.55 (0.22-10.94) | 0.66 |
| Overweight/Obesity | 23 | 3.04(1.29-7.11) | **0.01\*** | 2.25(0.78-6.53) | 0.14 |
| **Department** |  |  |  |  |  |
| Critical | 31 | 2.60 (1.23-5.51) | **0.012\*** | 5.24 (1.79-15.38) | **0.002\*** |
| **Years of clinical experience (years)** |  |  |  |  |  |
| >5 | 25 | 7.53 (2.78-20.32) | **<0.001\*** | 2.67 (0.58-12.39) | 0.209 |

**Age reference:** ≤30 years; **Type of hospital reference:** Public; **BMI reference:** Normal weight; **Department reference:** General; **Years of clinical experience:** ≤5 years
**\*p≤0.05**

## Discussion

A descriptive-analytical cross-sectional study design was used to assess the burden of work-related musculoskeletal disorders and associated factors among 118 Nurses working at Different hospitals in Sudurpaschim Province. The result of this study indicated a 47.46 percent prevalence of musculoskeletal disorders affecting any body part. However previous studies have documented various rates of musculoskeletal disorders over 12 months in a variety of populations.

The prevalence of work-related musculoskeletal disorders reported in the study is consistent with the prevalence rate reported by the studies in Thailand reported 12 months prevalence was 47.8% (23). Similarities in the results might be due to similar working cultures and techniques used while performing nursing procedures. The prevalence was slightly lower in India where 30.5% of health professionals reported having musculoskeletal symptoms than current study (10). The differences in the results might be due to the sampling population used, as the study in India included different categories of HCPs. However, the prevalence reported by this study is lower than the reported prevalence of musculoskeletal disorders in Dhaka city (77.6%), India (71%) (24,25). Unlike the result of the current study the prevalence of musculoskeletal disorders among nurses was significantly higher in Kaski, Nepal (86%), China (91.2%), Zimbabwe (95.7%), and Uganda (80.8%) (17,18,26). These differences in prevalence might be due to the study among the nurses of more advanced tertiary hospitals with a high flow of patients. And also might be due to a variety of disease conditions, practice settings, and the availability of basic clinical equipment.

Among the different body sites, the highest prevalence of musculoskeletal among nurses according to the study was at the Lower back (57.14%) followed by the ankle/feet (46.43%). This distribution pattern of musculoskeletal disorders among nurses is consistent with the study in Kaski Nepal with slightly higher prevalence of low back pain and ankle/foot pain 61.8 percent and 34.8 percent respectively (17). A similar sequence of affected body sites may result from similarities in nursing procedures and techniques. However, the slightly higher prevalence in Kaski could be attributed to the higher proportion of nurses from critical care units (17). Unlike the current study findings, many other studies documented different anatomical sites for higher prevalence i.e. in the neck among the nurses in Jordan and that can be explained by poor compliance to proper posture during working, and strenuous upper quadrant movements such as pulling or pushing (27)(27), in shoulder among the nursing assistants working in Honk Kong nursing homes, and which might be explained by variations in the types of work settings as lifting facilities in nursing homes might not be as adequate as in hospital settings (12). Previous studies have documented different rates of lower back pain over 12 months ranging between 40.0 to 80.0 percent. This study showed a 57.14 percent prevalence of low back pain. However, the study conducted in Kaski, Nepal, Dhulikhel Hospital and China showed a slightly higher prevalence of 61.8, 65.0 and 60.3 percent of low back pain among nurses respectively (17,28).

The difference in the result might be due to the greater number of respondents from the critical unit in the study of Kaski, Nepal, being a specific study for low back pain in the study of Dhulikhel, Nepal. Unlike the reported prevalence for low back pain in the current study, the study in India showed lower prevalences 52.0, 44.0, and 48.2 percent respectively in the study by (29,30). In contrast to the current study findings, a higher prevalence of low back pain was reported in a study conducted at Shahid Gangalal Hospital in Nepal (78.0%) by Adhikari and Dhakal in 2017, and in a study conducted in Malaysia (86.7%) by Krishnan et al. in 2021. The difference in prevalence could be attributed to the different work environments, as Gangal Hospital is a specialized cardiac treatment center, and researchers in the Malaysian study stated that the majority of the study participants were exposed to frequent high-risk and complex tasks (21,31). The study conducted in Mansoura University Children Hospital (MUCH) of Egypt strongly contradicts the findings of the current study as the most common site was the elbow (85.2%). This difference might be attributed to activities involving upper limb force in the manual handling and positioning of pediatric patients. The second most affected body site was the pelvis/ thigh with a prevalence of 74.9 percent and this can be explained by prolonged standing in patient care. The least body site for musculoskeletal disorders among nurses was the elbow in the current study. However, surprisingly the least site for pain was low back pain (37%) in Egyptian nurses (2).

The present study also concluded that slightly more than half (51.79%) of the nurses sought consultation and treatment in response to musculoskeletal disorders. However, only 37.5 percent of the nurses had sought any kind of treatment in Kaski, Nepal (17). This might be because the majority of nurses reported in Kaski changing the work setting which may relieve their symptoms. In the current study, only, 12.50 percent of the nurses reported changing work settings. However, the study in Kaski revealed the majority (89.5%) of the nurses had changed wards/department as a result of musculoskeletal disorders (17), and this can be explained by the different working environment of the medical college. The present study revealed that nearly half (46.43%) of the nurses had taken time off on leave as a result of musculoskeletal disorders.

However, a study in Zimbabwe showed 79.8 percent of nurses had to take a day off from work, which can be explained by a difference in nurse-patient ratio, as the total population of nurses at Zimbabwe’s central public hospital was 762 (32). Furthermore, more than half (48.21%) of the nurses in the current study reported participating in an exercise and posture program, which may have resulted in fewer sick days. Unlike this current study the finding of a study in Kaski, Nepal reported absenteeism (sick leave) as a consequence of musculoskeletal disorders was only 13.7 percent (17). This variation could be attributed to differences in work settings as well as the availability of services and benefits in their respective hospitals. Similarly, nurses may often tend to self-medication as a treatment for musculoskeletal pain to avoid taking sick leave.

This cross-sectional study also aimed to study nurse’s perception of job risk factors that may contribute to their musculoskeletal condition. According to the study findings, 69.64 percent of the nurses perceived that working in the same position for a long period was the most common job risk factor contributing to musculoskeletal disorders followed by reaching or working away from the body (67.86%) and work scheduling (64.29%). This finding is consistent with the previous reports indicating working in the same positions for long periods (29.8%) in Dhaka is the most important risk factor of musculoskeletal disorders (24). Similarly, studies in India, and Gangalal National Heart Centre, Nepal revealed prolonged standing as the most common (79% and 82%) risk factor of musculoskeletal disorders among nurses (30,31). The similarities in perception in all studies could be attributed to similar work settings, techniques, and procedures. Contradicting the finding of this study, 63.41 percent of nurses in Kaski, Nepal perceived that attending an excessive number of patients per day was the most common job risk factor contributing to musculoskeletal disorders followed by working in the same position for a prolonged period (56.2%) (17). Differences in the location of the hospital, cultural differences, and reporting of pain and disorders are adduced for the variation in perceived risk factors of musculoskeletal disorders.

Regarding the association of musculoskeletal disorders with socio-demographic characteristics of the respondents, the study findings revealed a statistically significant association between musculoskeletal disorders and age, and it is the most important predictor as musculoskeletal disorders were worse with an increase in age (OR= 6.92; CI =1.67-28.58). This finding is similar to the findings of the study in Kaski, Nepal in which age was significantly associated with musculoskeletal disorders (X^2^=4.15, p<0.04) (17). Likewise, the study findings are also consistent with the findings of a study conducted among Thai nurses (P<0.001, Unadjusted OR 1.70, 95% CI 1.15-2.28) and nurses in Uganda respectively (OR 1.03, 95% CI 1.01-1.06) (18).

Because age is frequently associated with job tenure, this association could be due to the cumulative effects of nurses’ long-term exposure to adverse working conditions that contribute to musculoskeletal disorders. The results of this study were in contrast with a study in Egypt where the prevalence of musculoskeletal complaints was statistically higher among nurses aging from 20 to 29 years compared to a higher age group (30-40 years) which can be referred to as the heavier manual tasks of younger nurses (2). As a result, it can be explained that, while the probability of MSDs increases with age, it may vary depending on the type of hospital and tasks performed.

The current study found no association between prevalence of musculoskeletal disorders, and marital status (p=0.22) and physical exercise (p=0.29). The results are in line with the study among nurses at Kaski Nepal, which also revealed no association between marital status (p=0.24) and physical exercise (p=0.82) (17).

The present study also concluded a statistically significant association between musculoskeletal disorders and Body mass index (X^2^=8.53; p<0.05). This finding is in line with several studies among nurses at Kaski, Nepal (X^2^=20.03; p<0.05) (17). Also, high BMI was the major predictor of musculoskeletal pain in the current study (Unadjusted OR 3.04; 95% CI 1.29-7.11). This finding is similar to a study among Thi nurses (23). It could be due to the contribution of the pressure of extra weight to the spine and muscles while performing their duties. The study showed a significant association between work experience and musculoskeletal disorders (OR=7.53; CI 2.78-20.32). This finding is similar to the study among nurses in Kaski, Nepal (X^2^=4.55, p<0.05). This association could be due to the cumulative effects of nurses’ long-term exposure to adverse working conditions that contribute to musculoskeletal disorders. The present study revealed a significant association between working area and musculoskeletal disorders (X^2^=6.39, p=0.03). However, the results contradict the findings of a study in Kaski where there was no significant association between musculoskeletal disorders and working areas, which might be attributed to treating the excessive number of patients in general wards, as addressed by the study (17).

In the current study, there were significant differences in reported musculoskeletal disorders across hospital type as musculoskeletal disorders were worse in the public hospitals as compared to the private hospital (OR 0.33, 95% CI 0.12-0.91; p <0.001). This finding is consistent with the study conducted in Uganda which showed musculoskeletal disorders were threefold worse in the public hospitals as compared to the private and private not-for-profit hospitals (OR 2.77, 95% CI 1.07-7.14) (18). The results could be attributed to the difference in human resources, working culture, and the adoption of advanced technology in both types of hospitals.

### Limitation

It is a cross-sectional study, therefore determining causality is difficult to navigate. Prospective studies are required to validate the retrospective analysis and its accuracy. Furthermore, the study did not examine the link between psychological factors and WMSDs. Further research into the implications of these risk variables on WMSDs is required.

## Conclusions

The current study revealed that work-related musculoskeletal disorders are common among nurses in Sudurpaschim province’s Different hospitals. This disorder had primarily affected the lower back, followed by the ankle and feet. To alleviate the symptoms of MSDs, most of the nurses reported seeking medical help followed by exercising and correcting posture. The perceived major risk factor for MSDs was working in the same position for a prolonged period. The findings showed a significant association of MSDs with the age and BMI of the nurses, type of hospital, working department, and work experience. The odds of having MSDs were high among nurses of age above 30 years, nurses with high BMI and work experience more than 5 years, and also high for those who worked in public hospital and critical care units.

## Data Availability

All relevant data are within the manuscript and its Supporting Information files.

## Acknowledgments

The authors would like to thank hospital authorities and the study participants for their time and views.

## Supporting Information

**S1 Fig. Problem Analysis Tree**

**S2 Fig. Ethical Approval**

**S1 File. Study Tool**

